# Quantitative MRI biomarker for classification of clinically significant prostate cancer: calibration for reproducibility across echo times

**DOI:** 10.1101/2024.01.25.24301789

**Authors:** Karoline Kallis, Christopher C. Conlin, Courtney Ollison, Michael E. Hahn, Rebecca Rakow-Penner, Anders M. Dale, Tyler M. Seibert

## Abstract

**Background:** Restriction Spectrum Imaging restriction score (RSIrs) is a quantitative biomarker for detecting clinically significant prostate cancer (csPCa). However, the quantitative value of the RSIrs is affected by imaging parameters such as echo time (TE).

**Purpose:** The purpose of the present study is to develop a calibration method to account for differences in echo times and facilitate use of RSIrs as a quantitative biomarker for the detection of csPCa.

**Methods:** This study included 197 consecutive patients who underwent MRI and biopsy examination; 97 were diagnosed with csPCa (grade group ≥ 2). RSI data were acquired three times during the same session: twice at minimum TE∼75ms and once at TE=90ms (TEmin_1_, TEmin_2_, and TE90, respectively). A proposed calibration method, trained on patients without csPCa, estimated a linear scaling factor (f) for each of the four diffusion compartments (C) of the RSI signal model. A linear regression model was determined to match C-maps of TE90 to the reference C-maps of TEmin_1_ within the interval ranging from 95^th^ to 99^th^ percentile of signal intensity within the prostate. RSIrs comparisons were made at 98^th^ percentile within each patient’s prostate.

We compared RSIrs from calibrated TE90 (RSIrs_TE90corr_) and uncorrected TE90 (RSIrs_TE90_) to RSIrs from reference TEmin_1_ (RSIrs_TEmin1_) and repeated TEmin_2_ (RSIrs_TEmin2_). Calibration performance was evaluated with sensitivity, specificity, area under the ROC curve, positive predicted value, negative predicted value, and F1-score.

**Results:** Scaling factors for C_1_, C_2_, C_3_, and C_4_ were estimated as 1.70, 1.38, 1.03, and 1.19, respectively. In non-csPCa cases, the 98^th^ percentile of RSIrs_TEmin2_ and RSIrs_TEmin1_ differed by 0.27±0.86SI (mean±standard deviation), whereas RSIrs_TE90_ differed from RSIrs_TEmin1_ by 1.81±1.20SI. After calibration, this bias was reduced to -0.41±1.20SI, representing a 78% reduction in absolute error. For patients with csPCa, the difference was 0.54±1.98SI between RSIrs_TEmin2_ and RSIrs_TEmin1_ and 2.28±2.06SI between RSIrs_TE90_ and RSIrs_TEmin1_. After calibration, the mean difference decreased to -0.86SI, a 38% reduction in absolute error. At the Youden index for patient-level classification of csPCa (8.94SI), RSIrs_TEmin1_ has a sensitivity of 66% and a specificity of 72%. Prior to calibration, RSIrs_TE90_ at the same threshold tended to over-diagnose benign cases (sensitivity 44%, specificity 88%). Post-calibration, RSIrs_TE90corr_ performs more similarly to the reference (sensitivity 71%, specificity 62%).

**Conclusion:** The proposed linear calibration method produces similar quantitative biomarker values for acquisitions with different TE, reducing TE-induced error by 78% and 38% for non-csPCa and csPCa, respectively.

## Introduction

Around 288,300 new cases of prostate cancer were expected in the United States in 2023 alone, accounting for nearly 29% of all cancer cases in men [1]. The standard procedure to diagnose clinically significant prostate cancer (csPCa) includes multiparametric MRI (mpMRI) prior to biopsy [2]. A biopsy is an expensive and invasive procedure, which has the potential for both overdiagnosis and underdiagnosis of csPCa, given that only a small part of the prostate gland [2] is sampled, which is why accurate imaging of the whole prostate gland is valuable. A large percentage of men suspected to have csPCa can be safely reassured without biopsy if the prostate gland appears normal on mpMRI, and further, when biopsy is needed, needles can be directed to the most suspicious lesions.

Diffusion-weighted imaging (DWI) plays a crucial role in mpMRI for the detection and characterization of csPCa [3]. Commonly, the apparent diffusion coefficient (ADC) is evaluated to identify csPCa. However, ADC is a simplification of the diffusion process, ignoring non-Gaussian restricted diffusion, and hence does not accurately represent the tumor properties [4]. More advanced DWI models have been designed to better represent the microstructure of real tissue. Examples include intravoxel incoherent motion imaging [5, 6], diffusion kurtosis imaging [7, 8], Vascular, Extracellular, and Restricted Diffusion for Cytometry in Tumor (VERDICT) [9–11], hybrid multidimensional MRI [12–15], and Restriction Spectrum Imaging (RSI) [9, 16].

In the case of RSI, the overall diffusion signal is represented as a weighted combination of signal contributions from multiple tissue compartments, each characterized by a different, fixed diffusion coefficient [16, 17]. Prior studies have developed and validated a four-compartment model for prostate cancer detection, with compartments corresponding broadly to restricted, hindered, and free diffusion, plus vascular flow [17–20]. The biomarker RSI restriction score (RSIrs) is derived by normalizing the signal from the model coefficient of the most restricted diffusion compartment (referred to as C_1_) by the median T_2_-weighted signal intensity within the prostate. RSIrs has proven to be a valuable biomarker for identifying csPCa [17–19], demonstrating superior detection of csPCa compared to ADC, and similar performance to that of PI-RADS v2.1 [18, 19]. However, acquisition parameters like echo time (TE) can influence the quantitative value of RSIrs [21].

To maximize the utility of RSIrs as a quantitative biomarker, we propose a simple calibration method, based on MRI biophysics, for data obtained at varying TE values. We demonstrate a partial linear relationship between RSIrs and TE and compare RSIrs values at two different TEs before and after calibration. By addressing the challenge of TE-dependent variability, we aim to advance the potential of mpMRI as a valuable clinical tool in the diagnosis and management of prostate cancer.

## Materials and Methods

### Patient Cohort

This study was conducted under the approval of the institutional review board at UC San Diego (IRB 210213) with a waiver of consent for prospective collection of RSI at multiple TEs. The research adhered to the principles outlined in the Declaration of Helsinki and all relevant regulations. 197 patients who underwent MRI examinations between 03/2021 and 02/2023 were included in the study.

MRI examinations were interpreted per routine clinical practice by ten board-certified and subspecialty fellowship-trained radiologists. 38 of the 197 scans were part of a separate research study for evaluation of treatment response and did not have an official clinical interpretation but did have proven high-grade csPCa on biopsy, with MRI-visible lesions defined on prior clinical scans by a board-certified radiologist (max 6 months prior) and confirmed on the research scan by a subspecialist radiation oncologist and prostate MRI scientist (10 years of experience). Segmentation of the whole prostate was performed using an FDA-cleared commercial AI tool (OnQ Prostate, Cortechs Labs, San Diego, CA, USA). Biopsy (systematic and targeted) and prostatectomy were conducted in accordance with clinical protocols, and both were examined by board-certified pathologists. Clinically significant prostate cancer (csPCa) was defined as grade group ≥ 2. In cases where patients underwent prostatectomy, the determination of the grade group was based on the final pathology report.

### MRI Acquisition

All MRI acquisitions were carried out on a 3T clinical GE scanner (Discovery MR750, GE Healthcare, Waukesha, WI, USA), using a 32-channel phased-array body coil encompassing the pelvis. The acquisition parameters can be found in **Table 1**. For all patients, three axial RSI scans were obtained that each sampled five b-values (0, 50, 800, 1500, 3000 s/mm^2^). Two of the scans were acquired with minimum echo time (TEmin_1_ and TEmin_2_, approximately 75ms), and the third series was acquired with an echo time (TE) of 90ms (TE90). A high-resolution T_2_-weighted reference image was also acquired with field of view (FOV) identical to the RSI volumes.

**Table 1:** Acquisition parameters for clinical multi-parametric MRI; RSI = Restriction Spectrum Imaging; T2W = T_2_ weighted MRI.

| Series | RSI - T<sub>E</sub>min | RSI – T<sub>E</sub>90 | T2W |
| --- | --- | --- | --- |
| FOV [mm] | 200*100 | 200*100 | 200*200 |
| Matrix (resampled dimensions) | 80*48 (128*128) | 80*48 (128*128) | 320*320 (512*512) |
| Number of Slices | 32 | 32 | 32 |
| Pixel size [mm] | 1.56*1.56 | 1.56*1.56 |  |
| Slice thickness [mm] | 3 | 3 | 3 |
| TR [ms] | 4500 | 4500 | 6176 |
| TE [ms] | 75.6-76.3 | 90 | 106 |
| b-values [s/mm<sup>2</sup>] (number of samples) | 0 (1), 50 (6), 800 (6), 1500 (12), 3000 (18) | 0 (1), 50 (6), 800 (6), 1500 (12), 3000 (18) | N/A |

Post-processing of the image data was performed using in-house software in MATLAB (version R2017a, MathWorks, Natick, MA, USA). DWI images were corrected for B_0_ inhomogeneity distortions, gradient nonlinearity, and eddy currents [20, 22, 23]. Multiple acquired DWI samples at specific b-values were averaged together and normalized by the median signal intensity of urine in the bladder at b = 0 s/mm^2^. RSI model fitting was performed as described in prior studies [17–19].

### RSIrs: Quantitative Biomarker

The RSI model is defined by the following formal:

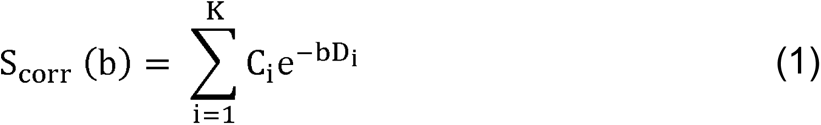

S_corr_ (b) defines the acquired averaged and noise-corrected DWI image at a particular b-value. D_i_ is the compartmental ADC. K denotes the number of tissue compartments. For this study, we modeled the signal using a four-compartment approach. C_i_ is the unit-less weighting factor describing the contribution of a particular compartment to the overall signal. The first compartment (C_1_) describes the signal from the slowest (intracellular restricted) compartment [17, 20].

The biomarker RSIrs is defined as C_1_, normalized by median signal intensity of the prostate at a b-value of 0 s/mm^2^ (mb0), i.e., the median T_2_-weighted signal of the whole prostate.

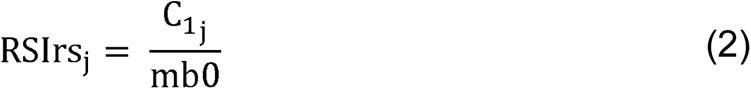

where j defines a voxel. Our emphasis was on high percentiles of RSIrs within the prostate, as the highest RSIrs values within each prostate are utilized for identifying csPCa on a patient level [19]. In this study, RSIrs comparisons were made at 98^th^ percentile within each patient’s prostate, as this is expected to be more stable than the maximum voxel and thus more robustly calibrated.

### Calibration Concept

Our hypothesis is that there exists a partial linear, echo-time dependent relationship among the acquired RSIrs-maps, as expressed in equation (3):

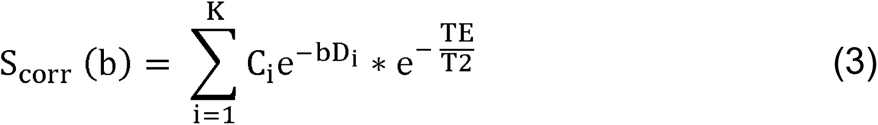

Where T_2_ defines the coefficient defining the T_2_-effect observed in the acquired images and TE the used echo time. As a demonstration of the concept, a linear regression model was optimized to partially fit RSIrs of TE90 to match RSIrs of TEmin_1_ within the range from the 95^th^ to the 99^th^ percentile of signal intensity within the prostate. By limiting the fitting to the range of high percentiles the influence of noisy data and imaging artifacts was minimized. Further, we focused on high percentiles of RSIrs because the highest values of RSIrs within each prostate are used to detect presence of csPCa on a patient level. As a result, linear scaling factors (f) were determined for each diffusion compartment (C), however currently only the information of C_1_ is included in RSIrs. Further, to ensure standardization, a calibrated mb0 value was determined for normalization purposes. The calibration of mb0 involved the artificial generation of DWI, utilizing the estimated scaling factors. This process effectively replicated the acquisition conditions with reference TE, see **Supplementary Figure 1**

Data from 100 control subjects (without csPCa) were used for training a partial linear regression model. The model was validated on the training set and a testing set comprised of 97 subjects with csPCa.

### Data Analysis

The 98^th^ percentile of RSIrs values within the whole prostate were compared between the reference TEmin_1_ (RSIrs_TEmin1_) scan and the TE90 (RSIrs_TE90_) scan, the TEmin_2_ (RSIrs_TEmin2_) scan, and the TE90 scan after calibration (RSIrs_TE90corr_). The difference between RSIrs_TEmin2_ and RSIrs_TEmin1_, acquired within minutes of each other with the same echo time, delineates the best achievable calibration and defines the error between various acquisitions with the same imaging parameters.

The mean and standard deviation of the differences in the 98^th^ percentile within each patient’s prostate between RSIrs_TEmin1_ and RSIrs_TE90_, RSIrs_TEmin2,_ and RSIrs_TE90corr_ were analyzed. A negative mean value describes that the quantitative value of the reference RSIrs_TEmin1_ acquisitions is higher than the corresponding RSIrs-map. A paired t-test of the 98^th^ percentiles between varying TE acquisitions was used to test for statistical significance (p<0.05).

Statistical performance of the calibration was investigated by comparing the sensitivity, specificity, positive predicted value (PPV), negative predicted value (NPV), and F1-score at a specific threshold (Youden index based on the ROC curve for RSIrs_TEmin1_). The area under the receiver operating characteristic (ROC) curve (AUC) was also reported.

## Results

218 consecutive patients who underwent MRI examinations between 03/2021 and 02/2023 were included in the study. Patients were excluded from the study if they had undergone prior treatment for prostate cancer, had a hip implant, or had a PIRADS score greater than 1 and no available biopsy result performed within 182 days of MRI acquisition. Further, patients were excluded because one of the TE acquisitions was missing, the imaging protocol did not match the norm for this analysis, or the pathology report was inconclusive. In total, 197 patients were included in the study. 97 of the 197 patients were identified to have csPCa, while 100 had only benign tissue or grade group 1 cancer. Further details of the patient cohort are presented in **Table 2**.

**Table 2:** Patient Characteristics range IQR= Inter quartile range, MRI = magnet resonance imaging; PSA = prostate-specific antigen.

| Parameter | Specification | Value |
| --- | --- | --- |
| Number of patients | Total | 197 |
| #patients with Urolift | Total | 3 |
| Recruiting Time Frame | Range | 03/2021 – 02/2023 |
| Age [a] | Median (IQR) | 69 (10) |
| Time from MRI to biopsy [d] | Median (IQR) | 56 (72) |
| PSA at time of MRI [ng/ml] | Median (IQR) | 6.0(5.1) |
| Prostate volume [ml] | Median (IQR) | 49 (37) |
| PSA density [ng/ml<sup>2</sup>] | Median (IQR) | 0.11 (0.11) |
| Biopsy naïve | Yes | 44 |
|  | No | 44 |
|  | Unknown | 109 |
| Best available pathology | Systematic only | 73 |
|  | Targeted only | 2 |
|  | Systematic + Targeted | 76 |
|  | No Biopsy | 37 |
|  | Unknown Biopsy type | 9 |
|  | Prostatectomy | 23 |
| PI-RADS Score | 1 | 68 |
|  | 2 | 10 |
|  | 3 | 18 |
|  | 4 | 34 |
|  | 5 | 29 |
|  | No Score | 38 |
| Gleason Grade Group | Benign | 72 |
|  | 1 | 28 |
|  | 2 | 41* |
|  | 3 | 31* |
|  | 4 | 8* |
|  | 5 | 17* |

\*clinically significant prostate cancer

Scaling factors for C_1_, C_2_, C_3_ and C_4_-maps were estimated at 1.70, 1.38, 1.03, and 1.19, respectively, for converting TE90 data to the TEmin reference. Examples illustrating cases are presented in **Figure 1** (absence of csPCa) and **Figure 2** (presence of csPCa).

**Figure 1:**
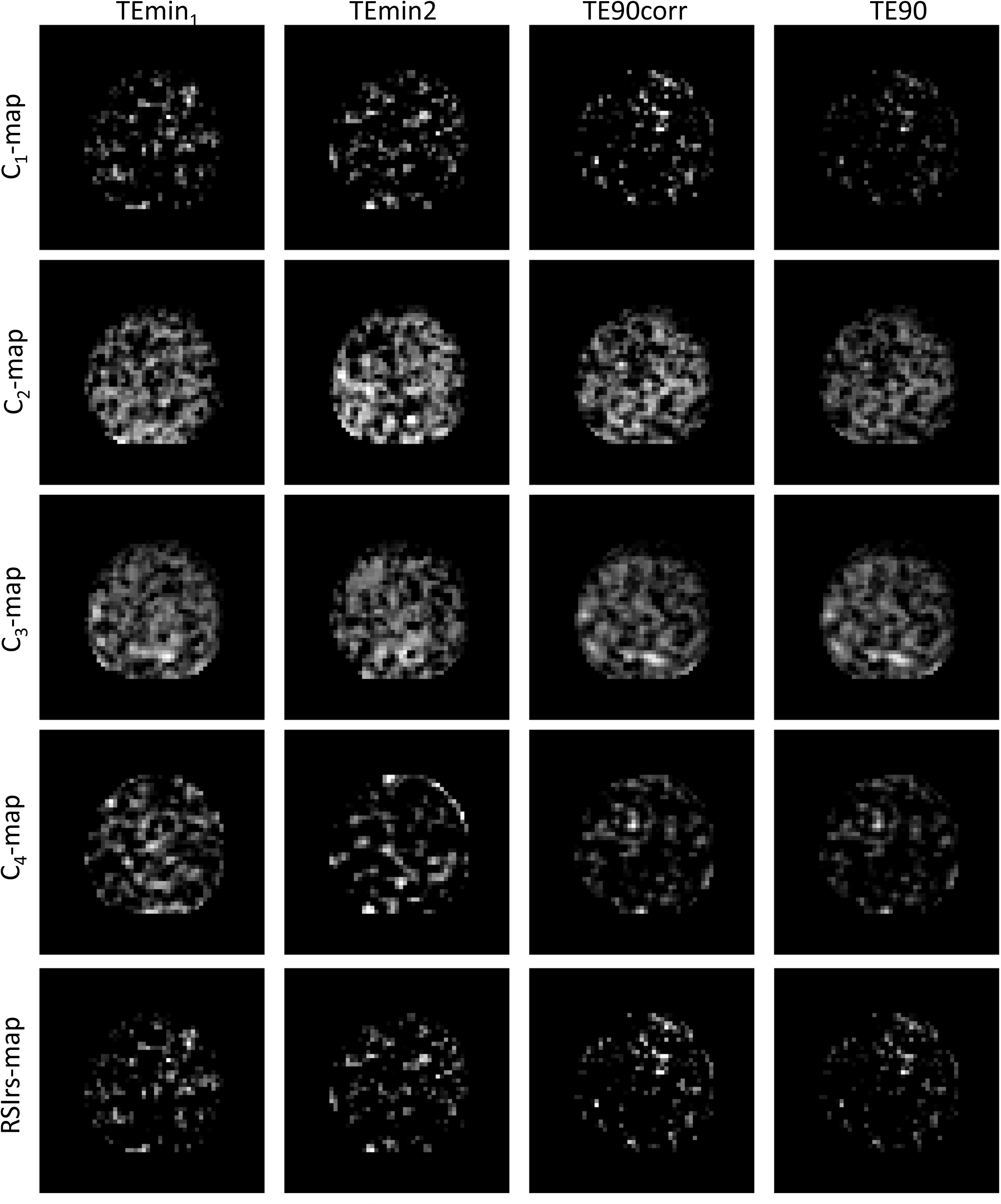
C-maps and RSIrs-maps for a patient without csPCa. Images are shown for the TEmin_1_, TEmin_2_, TE90, and TE90corr acquisitions.

**Figure 2:**
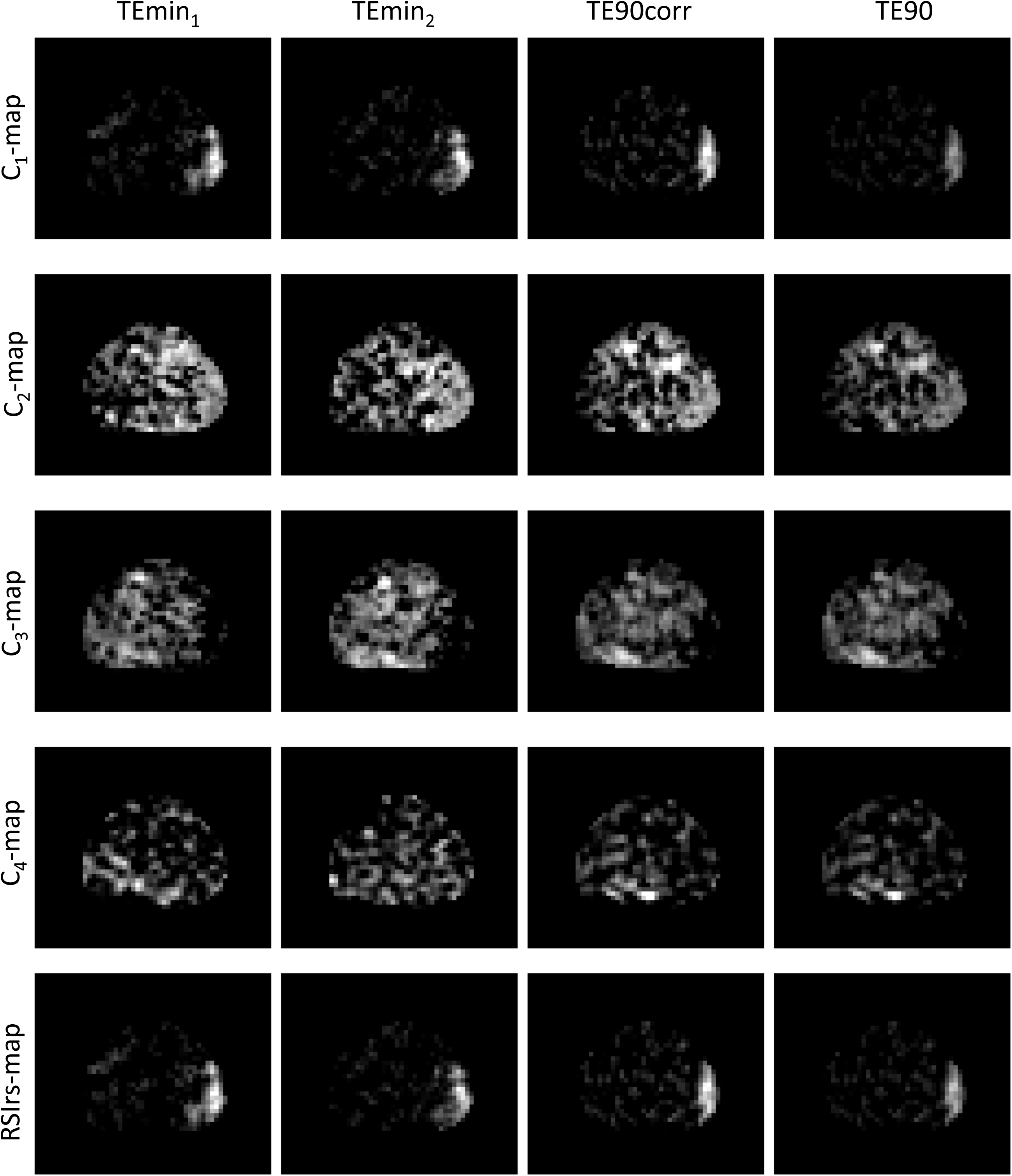
C-maps and RSIrs-maps for a patient with csPCa (PIRADS 5 lesion). Images are shown for the TEmin_1_, TEmin_2_, TE90, and TE90corr acquisitions.

Figure 3 illustrates the 98th percentile of RSIrs_TE90_, RSIrs_TE90corr_, and RSIrs_TEmin2_ within the prostate for each patient, comparing them to the reference (98th percentile of RSIrs_TEmin1_). In non-csPCa cases, a difference of 0.27±0.86 SI (p<0.01) was observed between the 98^th^ percentiles of RSIrs_TEmin2_ and RSIrs_TEmin1_. The difference between RSIrs_TE90_ and RSIrs_TEmin1_ was 1.81±1.20 SI (p<0.01), indicating that a ∼15 ms change in TE led to a 7-fold increase in the difference between RSIrs measurements, compared to a repeat acquisition at the same TE. Following calibration, however, the bias between the two series was reduced to -0.41 SI (p<0.01), representing a 78% reduction in absolute error. In patients with csPCa, the 98^th^ percentile of RSIrs differed by 0.54±1.98 SI (p<0.01) between RSIrs_TEmin2_ and RSIrs_TEmin1_. Prior to calibration, the disparity between 98th percentile of RSIrs_TE90_ and RSIrs_TEmin1_ was 2.28±2.06 SI (p<0.01), more than 4 times larger than the difference between repeated acquisitions at the same TE. Following calibration, this average difference improved to -0.86 SI (p<0.01), signifying a 38% reduction in absolute error.

**Figure 3:**
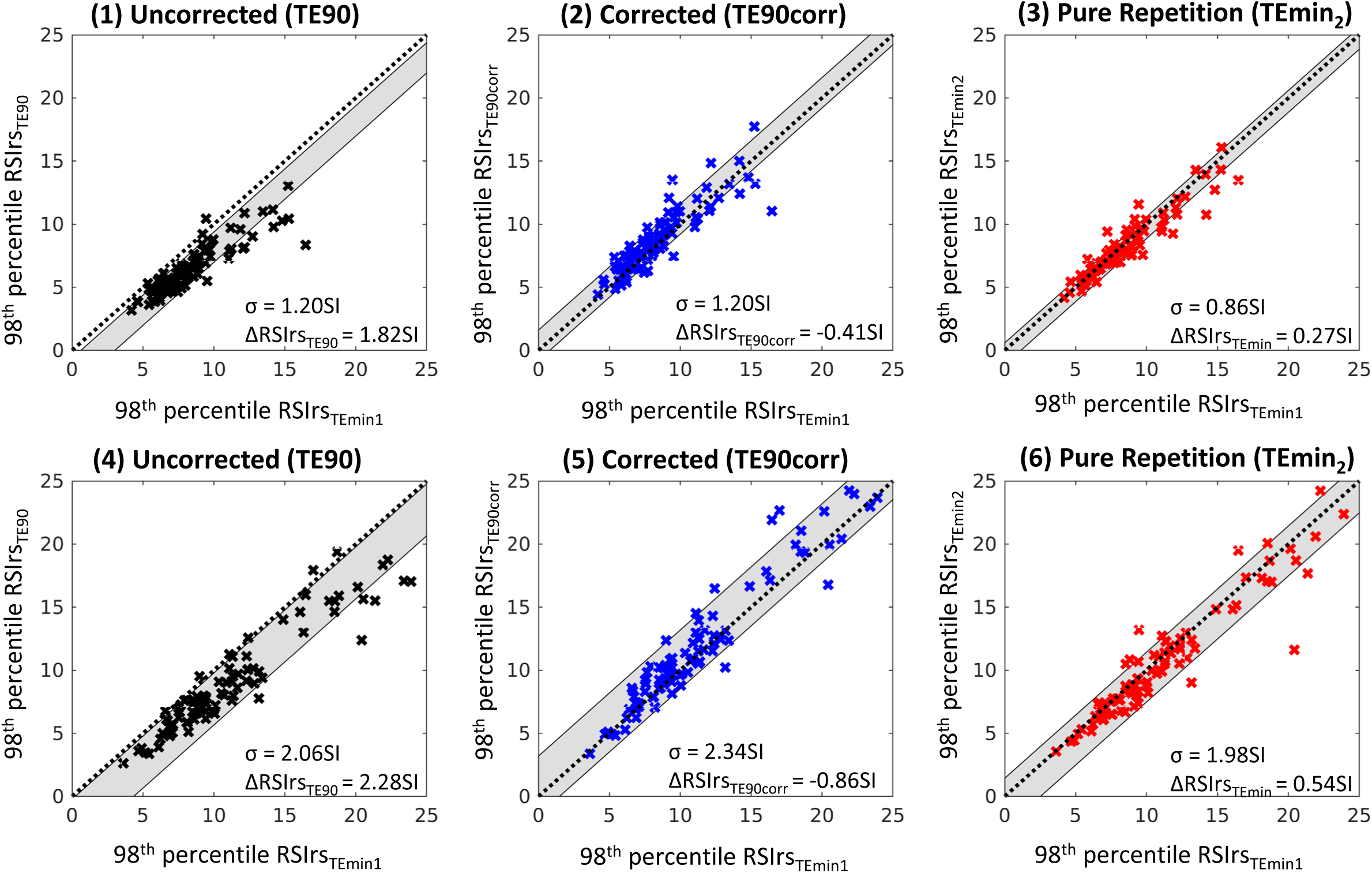
Comparison of the 98th percentile of RSIrs_TEmin1_ to that of RSIrs_TE90_ for benign cases (1-3) and csPCa (4-6) cases, for the RSIrs_TE90_ (1,4), RSIrs_TE90corr_ (2,5) and RSIrs_Temin2_ (3,6) acquisitions. Standard deviation (indicated by σ as well as gray color wash) and mean difference of the reference, 98th percentile of RSIrs_TEmin1_, to the 98th percentile of RSIrs_TE90_, RSIrs_TEmin2_ and RSIrs_TE90corr_ ( ΔRSIrs) indicating model bias. B lack dashed lines indicate hypothetical perfect relation.

**Supplementary Figure 2** presents the 98th percentile of RSIrs_TEmin1_, RSIrs_TEmin1_corr_, and RSIrs_TEmin2_ within the prostate for each patient, comparing them to the reference (98th percentile of RSIrs_TE90_), when TE90 is utilized as the reference sequence for calibration instead of TEmin1. The threshold defined by the Youden index for the classification of csPCa was determined to be 8.94SI. RSIrs_TEmin1_ has a sensitivity of 66% and a specificity of 72%. Prior to calibration, RSIrs_TE90_ exhibits a low sensitivity (44%) coupled with high specificity (88%). Post-calibration, RSIrs_TE90corr_ performs more similarly to the reference (sensitivity 71%, specificity 62%), see **Table 3** and **Supplementary Figure 3**.

**Table 3:** Comparison of statistical parameters for the detection of csPCa at threshold defined through Youden index of 98^th^ percentile of RSIrs.

| Parameter | RSIs |  |  |  |
| --- | --- | --- | --- | --- |
|  | TE90 | TE90corr | TEmin1 | TEmin2 |
| Sensitivity | 44% | 71% | 66% | 57% |
| Specificity | 88% | 62% | 72% | 72% |
| PPV | 78% | 64% | 70% | 66% |
| NPV | 62% | 69% | 69% | 63% |
| AUC | 0.73 | 0.72 | 0.72 | 0.71 |
| F1-score | 56% | 68% | 68% | 61% |

## Discussion

We present a straightforward, physics-based, method for calibration between different echo times. The calibration method demonstrated an improvement of inherent bias between RSIrs_TE90_ and RSIrs_TEmin1_. Residual error (in 98th percentile of RSIrs) after calibration was 78% percent smaller in prostates without csPCa and 38% smaller in prostates with csPCa.

Overall, calibration improved the similarity of the statistical performance for the detection of csPCa of RSIrs_TE90_. The difference in sensitivity and specificity decreased from 22% to 5% and 16% to 10%, with an identical F1-score between RSIrs_TEmin1_ and RSIrs_TE90corr_ of 68%.

A significant difference between RSIrs_TEmin1_ and RSIrs_TEmin2_ was observed in two acquisitions with identical imaging parameters acquired only a few minutes after each other without the patient leaving the scanner. Possible explanations for these changes could be explained by patient motion, changes in rectal gas, and hardware factors like pre-scan signal normalization or gradient heating. These factors add complexity to the calibration process but also define the limits of achievable correspondence between scans. The absolute differences between the 98^th^ percentile of RSIrs_TEmin1_ and RSIrs_TEmin2_ were larger for grade group 4 and 5, as shown in **Supplementary Figure 3**. The presented numbers reflect increased uncertainty in higher grade groups, making calibration for csPCa more challenging.

Due to the impact of noise in low percentiles and artifacts in high percentiles, concentrating on the 95^th^ to 99^th^ percentile is reasonable for the clinical use case of detection of csPCa, where the highest RSIrs values in a patient’s prostate are known to drive quantitative performance of csPCa detection at the patient level [18, 19]. We note, though, that a focus on calibrating high values may imply relatively poorer calibration in voxels with lower values. The clinical utility of values with near-zero values is unclear, so this may not be consequential in practice. Moreover, prostate images after calibration suggest the proposed method improves consistency with reference images (Figure 1, Figure 2).

An important limitation of this study is that the method solely addresses variations in echo time. Another limitation of the present work is reliance on data from a single institution and scanner manufacturer. To establish calibration across scanners from different manufacturers and accommodate changes in imaging parameters such as field strength and b-values, more sophisticated techniques like histogram matching [24, 25] or the incorporation of machine learning methodologies [26] would be necessary. The present study is instructive, though, as it reveals that even minor variations of 15ms in TE can result in significant differences in quantitative measurements that require careful calibration and demonstrates physics-based correction for these differences. Our findings may support protocol standardization, as much as possible, in the application of quantitative diffusion MRI to better ensure accurate and reliable results.

In conclusion, this study showed the feasibility of a straightforward calibration method to account for echo time differences for images acquired at the same scanner. DWI metrics are highly sensitive to changes in TE. A change of ∼15ms in TE resulted in errors 5-fold (csPCa cases) or 10-fold (benign prostates), compared to the errors incurred by simply repeating the acquisition with a consistent TE. The implementation of a simple linear calibration proves effective in generating comparable quantitative biomarker values across acquisitions with differing TE, resulting in a substantial reduction of TE-induced errors by 44% and 78% for csPCa and benign prostates, respectively.

## Data Availability

The datasets generated during and/or analyzed during the current study are not publicly available due to required IRB approval but are available from the corresponding author on reasonable request.

