## Supplemental Material for "Quantitative MRI biomarker for classification of clinically significant prostate cancer: calibration for reproducibility across echo times"

**Supplemental Materials**

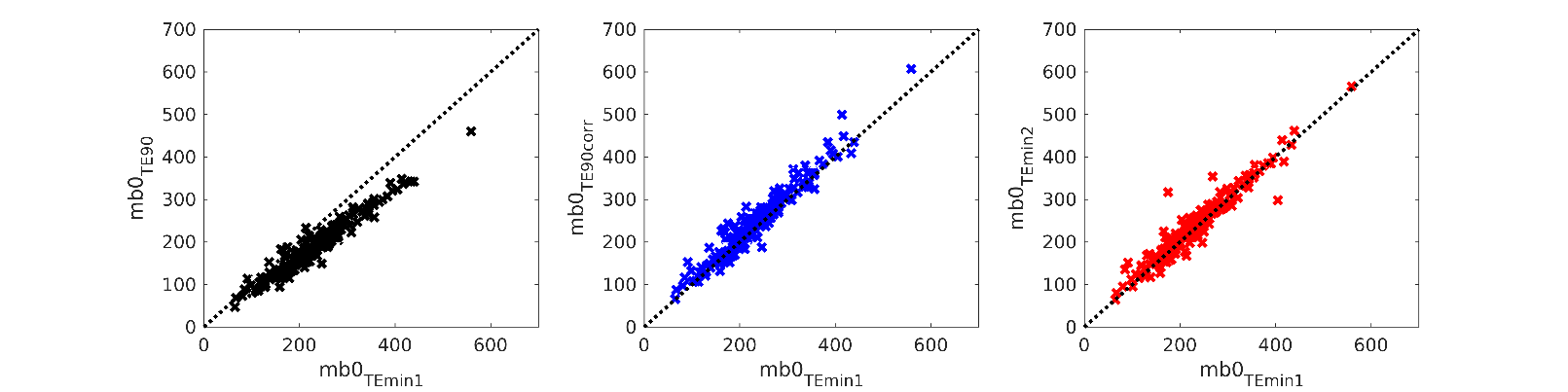
**Supplementary Figure 1:** Comparison of mb0 TEmin_1_ for all cases to mb0 TE90 , mb0 TE90corr and mb0 TEmin_2_ . Black dashed lines indicate hypothetical perfect relation.

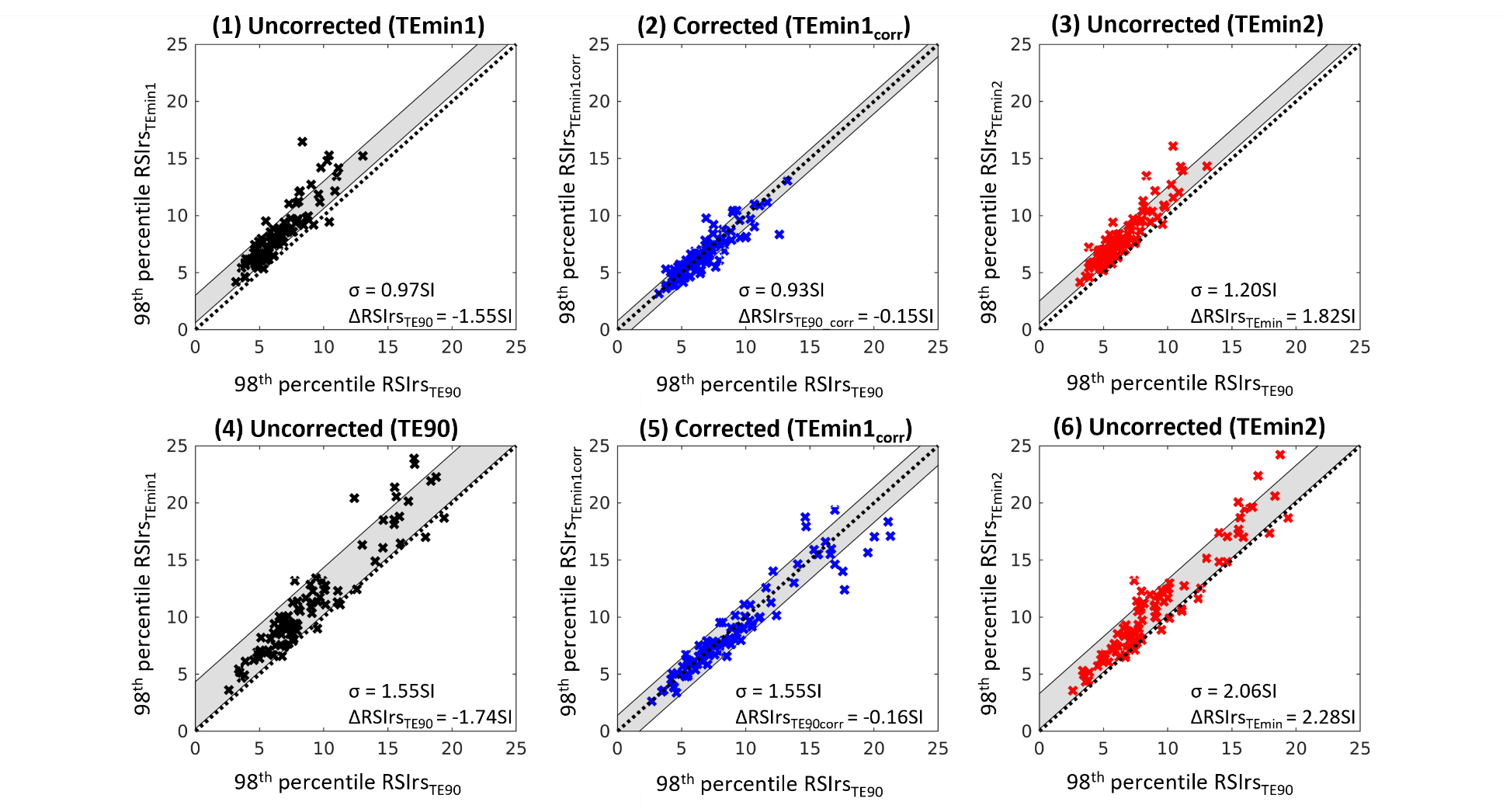

**Supplementary Figure 2:** Comparison of the 98th percentile of RSIrs_TE90_ for benign cases (1-3) and csPCa (4-6) cases within the prostate to the 98th percentile of RSIrs_TEmin1_ (1,4), RSIrs_TE90corr_ (2,5) and RSIrs_TEmin2_ (3,6). Standard deviation (indicated by σ as well as gray color wash) and mean difference of the reference, 98th percentile of RSIrs_TE90_, to the 98th percentile of RSIrs_TEmin1_, RSIrs_TEmin2_ and RSIrs_TE90cor_r (ΔRSIrs) indicating model bias. Black dashed lines indicate hypothetical perfect relation.

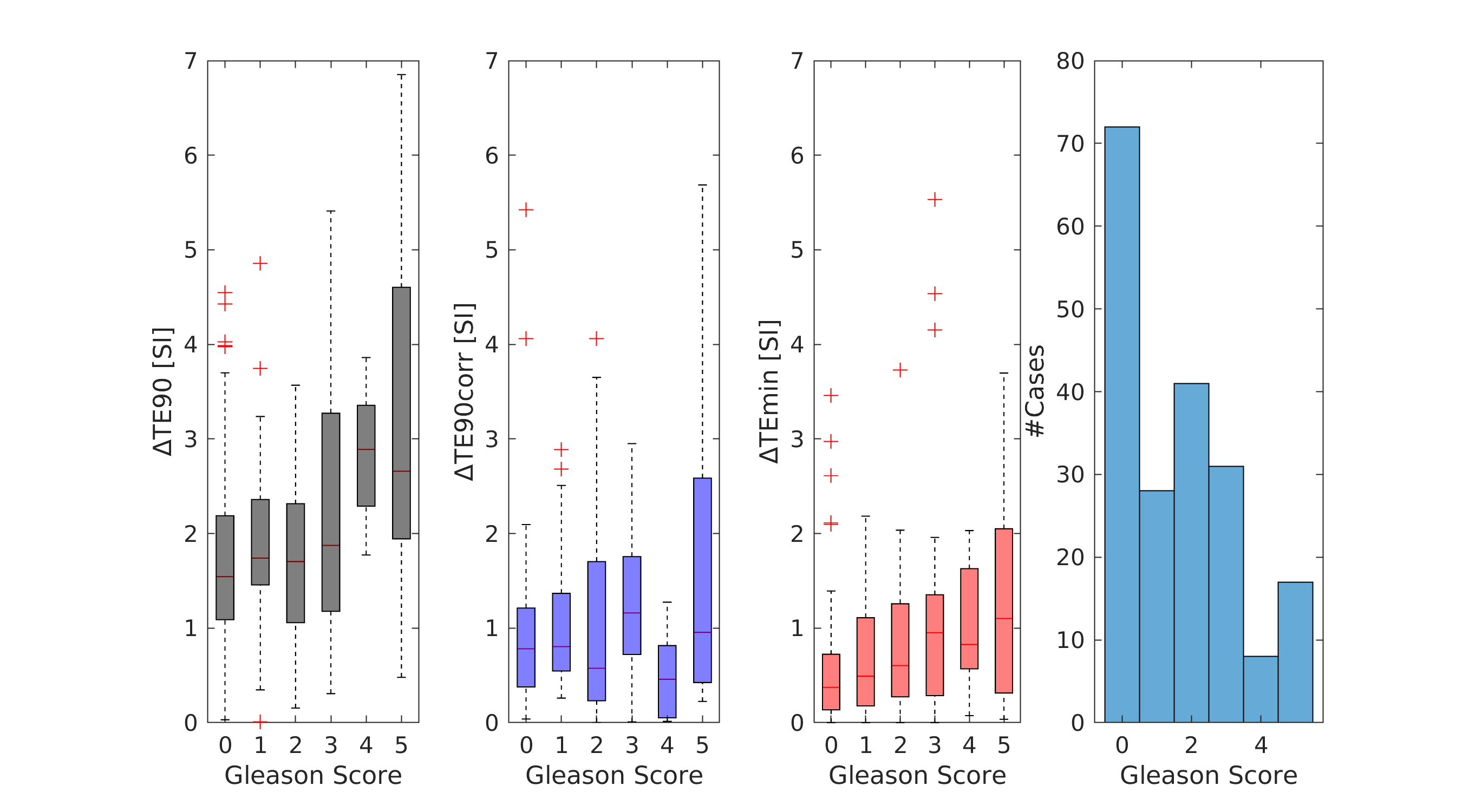

**Supplementary Figure 3:** Absolute differences between TE90, TE90corr, and TEmin2 and the reference acquisition TEmin1 for each Gleason grade group. Gleason Score =0 indicates benign cases. The histogram illustrates the frequency distributions of Gleason scores in the patient cohort.

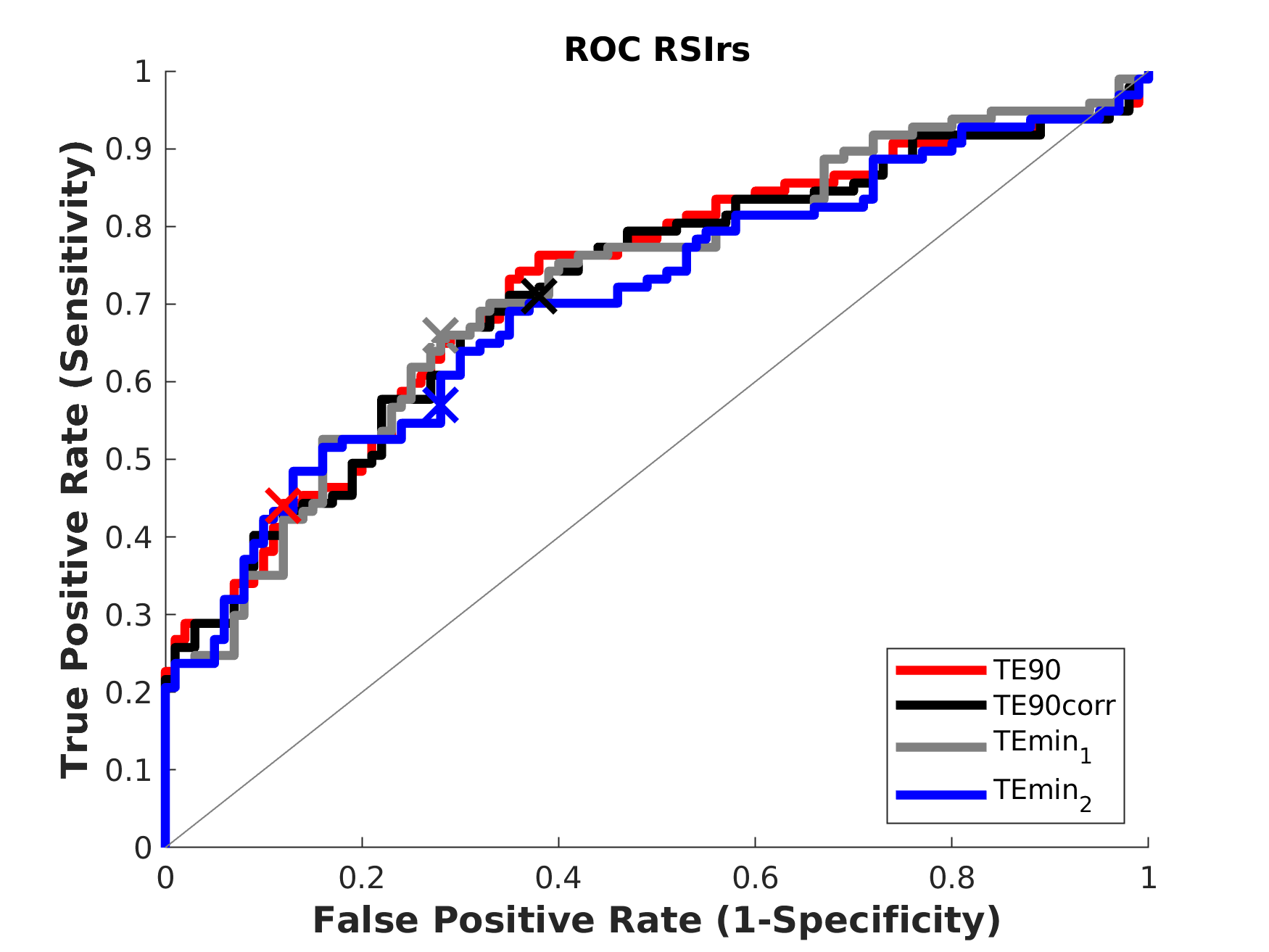

**Supplementary Figure 4:** ROC curve for classification of csPCa using the 98^th^ percentile of RSIrs within the prostate as the biomarker. The grey line indicates a model performance better than randomness. Crosses indicate the classification performance at the threshold defined by Youden index.
